# Accuracy and reliability of self-administered visual acuity tests: Systematic review of pragmatic trials

**DOI:** 10.1101/2023.02.03.23285417

**Authors:** Arun James Thirunavukarasu, Refaat Hassan, Aaron Limonard, Shalom Vitreous Savant

**Author notes:** **Corresponding author:** Arun Thirunavukarasu, Corpus Christi College, Cambridge, CB2 1RH, UK. **Author contributions:** AJT conceived and led the study. AJT prepared the study protocol and implemented the search strategy. AL, SVS, and AJT undertook abstract screening; AJT and RH undertook full-text screening, quality assessment, and data extraction. AJT conducted data analysis and produced figures. AJT drafted the manuscript with feedback from AL, SVS, and RH. All authors approved the final draft for submission.

## Abstract

**Background:** Remote self-administered visual acuity (VA) tests have the potential to allow patients and non-specialists to assess vision without eye health professional input. Validation in pragmatic trials is necessary to demonstrate the accuracy and reliability of tests in relevant settings to justify deployment.

**Methods:** A systematic review was undertaken in accordance with a preregistered protocol (CRD42022385045). The Cochrane Library, Embase, MEDLINE, and Scopus were searched. Screening was conducted according to the following criteria: (1) English languge; (2) primary research article; (3) visual acuity test conducted remotely; (4) no clinical administration of remote test; (5) accuracy or reliability of remote test analysed. There were no restrictions on trial participants. Quality assessment was conducted with QUADAS-2.

**Results:** Of 1227 identified reports, 10 studies were ultimately included. One study was at high risk of bias and two studies exhibited concerning features of bias; all studies were applicable. Three trials—of DigiVis, iSight Professional, and Peek Acuity—from two studies suggested that accuracy of the remote tests was comparable to clinical assessment. All other trials exhibited inferior accuracy, including conflicting results from a pooled study of iSight Professional and Peek Acuity. Two studies evaluated test-retest agreement—one trial provided evidence that DigiVis is as reliable as clinical assessment. The three most accurate tests required access to digital devices. Reporting was inconsistent and often incomplete, particularly with regards to describing methods and conducting statistical analysis.

**Conclusions:** Remote self-administered VA tests appear promising, but further pragmatic trials are indicated to justify deployment at scale to facilitate patient or non-specialist led assessment which could augment teleophthalmology, non-specialist eye assessment, pre-consultation triage, and autonomous long-term monitoring of vision.

## Introduction

Visual acuity (VA) is a measure of the functional resolution of vision, and is assessed before every ophthalmological, optometric, and orthoptic examination to inform decision making. Generally, VA assessment involves a clinician appraising the smallest optotype the patient can read while at a standard distance away from an illuminated chart. Self-administered VA tests provide patients with a means of monitoring their vision without having to be examined by an eye health professional. These tests may augment telehealth services, as VA assessment is an integral part of any eye examination. Adoption of self-administered VA tests may reduce the burden on strained ophthalmology resources by enabling non-specialists to triage with knowledge of visual function; by improving referral quality with provision of VA data; and by facilitating autonomous monitoring of vision by patients with chronic eye conditions (who otherwise require frequent clinic appointments).^1–3^ Many remote visual acuity tests have been developed, but most require administration in real time by a trained clinician, as required with conventional VA assessment (such as with Snellen or ETDRS chart).^4–6^

As the requirement for clinical examination limits the usefulness of ophthalmic telehealth services, platforms facilitating further examination without physical attendance will serve as important components of any improved suite for remote consultation.^3,6^ Many remote clinician-administered tests have been developed and applied in a wide variety of clinical settings around the world.^1,4^ However, removing the requirement for clinical administration is essential for remote VA tests to empower non-specialists and patients to test their vision. In recent years, newer platforms have emerged with this capability,^7^ and impetus for validation and implementation has been provided by the COVID pandemic.^1,8,9^ Most validation studies either involve clinical administration or are conducted in optimised clinical environments, unrepresentative of remote self-testing by patients at home or away from the eye unit.^4^ Pragmatic trials are essential to demonstrate that remote tests are useful for generating actionable VA data without skilled supervision—artificial environments are expected to inflate accuracy and reliability.^10,11^ Validation data generated in unrealistic settings provides weaker justification for subsequent clinical deployment than results generated in real-world conditions.^11^

Here, a systematic review was undertaken to identify remote self-administered VA tests; appraise the quality of their validation data; and compare these tests to conventional visual acuity testing. Specifically, the accuracy and reliability of VA self-tests were gauged, to establish the clinical utility of available platforms. All trials were pragmatic in that remote tests were administered without real-time clinical input, away from artificially ideal conditions. This evidence synthesis serves as a point of reference for clinicians, patients, and policy makers interested in identifying appropriate platforms to facilitate visual acuity assessment without requiring eye health service involvement.

## Materials and Methods

This systematic review adhered to PRISMA guidance, according to a prospectively registered protocol on PROPERO (identifier CRD42022385045). On 23 December 2022, The Cochrane Library, Embase (via OVID), MEDLINE (via PubMed), and Scopus were searched for the following: (“visual acuity”) AND (“remot*” OR “portable” OR “home based”) AND (“test” OR “assessment” OR “examination”). Previously published reviews were also searched for relevant studies.^4–7^ Duplicates were removed by a single researcher using Zotero (version 6.0.19-beta.15+6374aea1c; Digital Scholar, Vienna, Virginia, USA). Abstract and full text screening were undertaken by two independent researchers in Rayyan,^12^ with a third researcher acting as arbiter to resolve disagreement. The following inclusion criteria were employed: (1) Written in the English language; (2) Is a peer-reviewed primary research article; (3) Study examines a visual acuity test undertaken out of clinic (*i*.*e*. remotely); (4) The remote test does not require a clinically trained administrator (*i*.*e*. patient-led); (5) The remote patient-led test is compared to clinical or repeated remote visual acuity measurements to assess accuracy or reliability, respectively. No restrictions were placed on participant characteristics or test modality.

Risk of bias and concerns regarding applicability were appraised with the QUADAS-2 framework by a single researcher, with a second research verifying each appraisal.^13^ One researcher undertook data extraction for each included study, with a second independent researcher verifying every entry. Data gathered included details about participants, index tests, reference tests, measured outcomes, and study designs; and for index test-retest reliability and accuracy (*i*.*e*. comparison to clinical reference test), the bias and limits of agreement of Bland-Altman plots, correlation coefficients and *p* value, and *t*-test *p* value. For consistency, bias was expressed as the mean difference between reference and index test, such that positive values indicated that the reference test tended to provide a higher value (*i*.*e*. where the index test overestimated visual acuity). Where studies provided individual participants’ VA data without further analysis, the two-way random effects intraclass correlation coefficient (ICC) was calculated, and unpaired two-samples *t*-test was conducted. For studies exhibiting Bland-Altman plots without reporting figures for the bias and limits of agreement, manual interpolation was conducted with WebPlotDigitizer (version 4.6.0; Ankit Rohatgi, Pacifica, California, USA; company). Meta-analysis was planned but ultimately precluded by a lack of trials testing the same platform. Data extraction and quality assessment were conducted in in Microsoft Excel for Mac (version 16.57; Microsoft Corporation, Redmond, Washington, USA). Data analysis was conducted in R (version 4.1.2; R Foundation for Statistical Computing, Vienna, Austria).^14–16^ Tables were produced in Microsoft Excel for Mac. Figures were produced in R and modified with Affinity Designer (version 1.10.4; Pantone LLC, Carlstadt, New Jersey, USA).

## Results

The undertaken literature search and screening process is summarised in Figure 1. 10 studies were included from 1227 identified reports.^17–26^ Fulfilling criterion (3) necessitated that trials were pragmatic in that remote tests were conducted out of the eye clinic.^27^ Hyperacuity tests and survey-based self-assessment were excluded.^28–31^ To fulfil criterion (4), tests had to be patient-led: while tests administered by parents for paediatric patients were acceptable, involvement of clinicians or other trained personnel justified exclusion.^32–34^ Criterion 5 mandated exclusion of studies involving tests which did not provide visual acuity measurements which could be compared to conventional clinical assessment or repeated remote measurement.^35–37^

**Figure 1.**
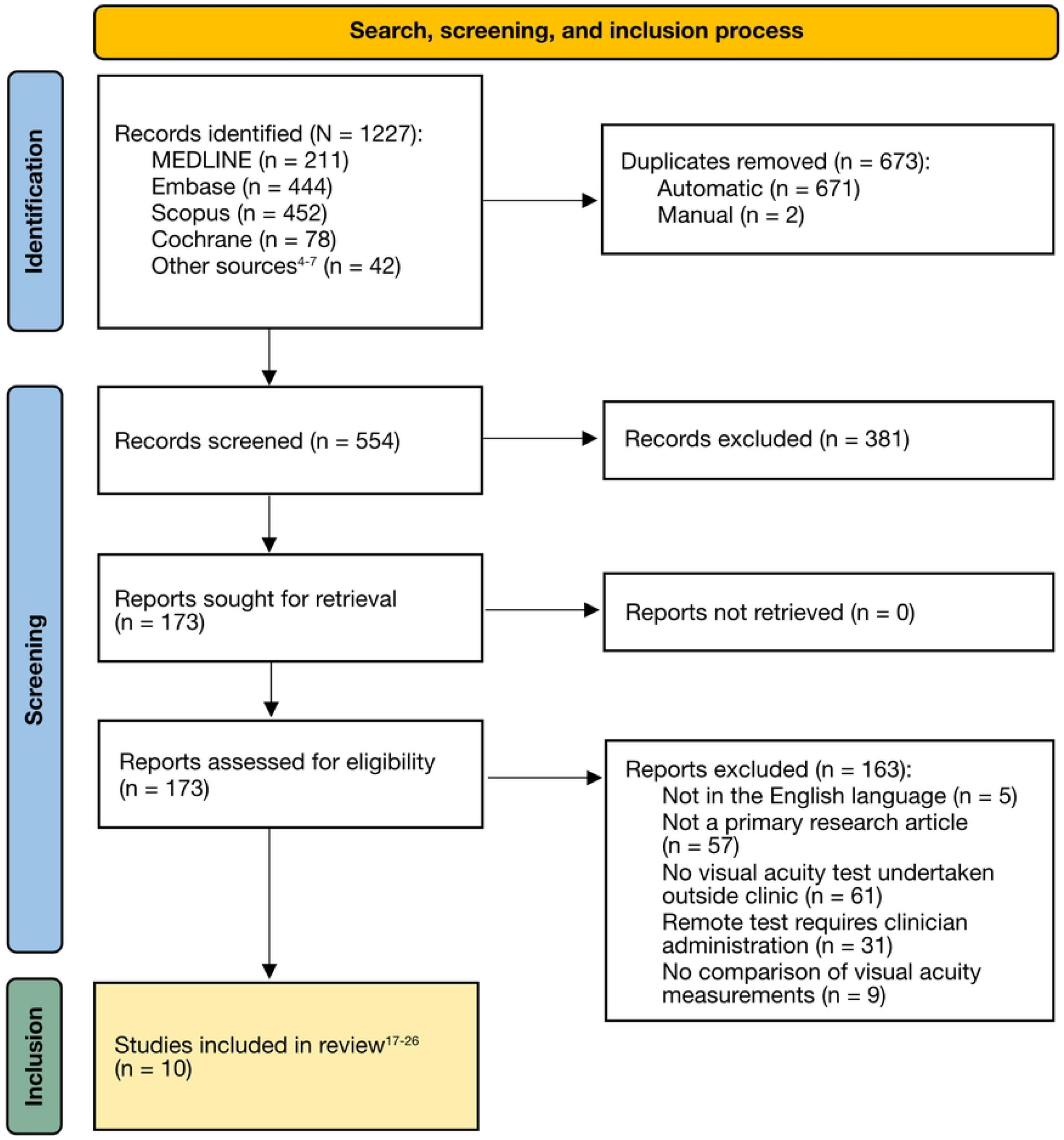
PRISMA flowchart. Illustrating the literature search, screening process, and articles included in this review. PRISMA = Preferred Reporting Items for Systematic Reviews and Meta Analyses; MEDLINE = Medical Literature Analysis and Retrieval System Online.

Study characteristics are summarised in Table 1. Most studies were prospective cross-sectional surveys, with just one retrospective case-control study. 6 of 10 studies reported conflicts of interest, suggesting that many validation studies were not undertaken by research teams independent from the trialled product—a potential source of reporting bias. However, none of the included studies received private funding, such as from product manufacturers. The number of participants ranged from 7 to 148 (median = 50.5). Reported participant age ranged from 3 to 95 years old—spanning most of the paediatric and adult ophthalmology case load.

**Table 1.**
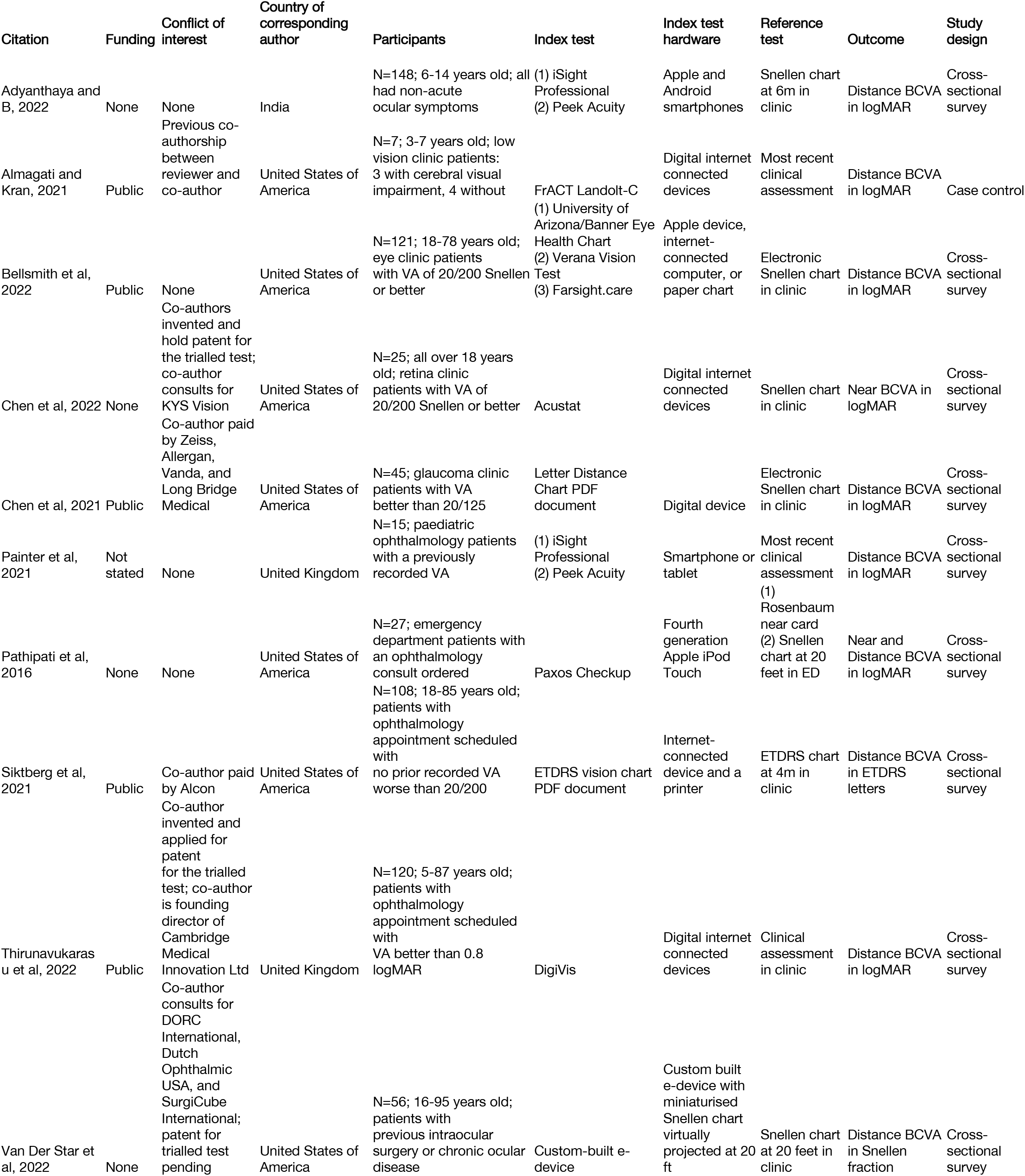
Characteristics of each of the included studies. BCVA = best corrected visual acuity; VA = visual acuity; logMAR = logarithm of the minimum angle of resolution; PDF = portable document format; ETDRS = Early Treatment of Diabetic Retinopathy Study.

Most trialled tests required access to digital devices: exceptions required a paper chart or custom-built e-device; both provided by the investigators.^19,26^ One study required patients to print a physical chart sent to their digital device.^24^ Risk of bias judged with QUADAS-2 was generally low, as illustrated in Figure 2 and S1 Figure. No major concerns regarding applicability were highlighted during QUADAS-2 appraisal, likely due to stringent inclusion criteria ensuring all studies applied patient-led tests remotely.

**Figure 2.**
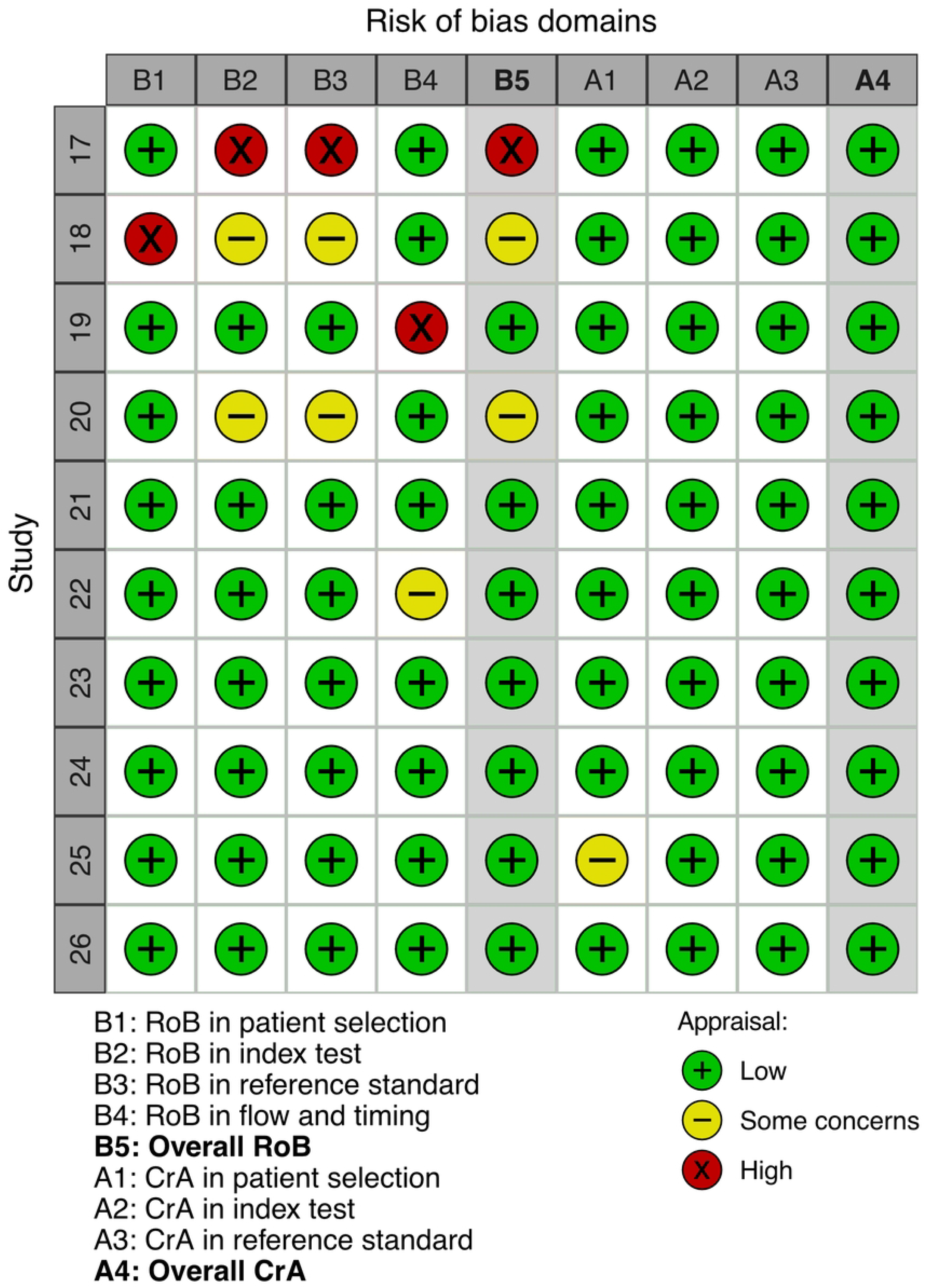
Risk of bias and inapplicability appraisals for each included study. Appraised with the QUADAS-2 framework. QUADAS-2 = Quality Assessment of Diagnostic Accuracy Studies 2; RoB = risk of bias; CrA = concerns regarding applicability.

All studies gauged accuracy by comparing remote measurements to assessment in clinic (Table 2). The reference test was not consistently defined in three studies,^18,22,25^ and Snellen chart was used in four studies;^19–21,23,26^ as opposed to the gold-standard Early Treatment for Diabetic Retinopathy Study (ETDRS) chart which was used consistently in just one study.^24^ One study trialling FrACT provided individualised data which enabled calculation of the bias and intraclass correlation coefficient, but its small sample size and retrospective design were discussed by the authors as significant limitations necessitating further validation.^18^ One trial of a custom e-device did not report any statistical analysis or individual data.^26^

**Table 2.**
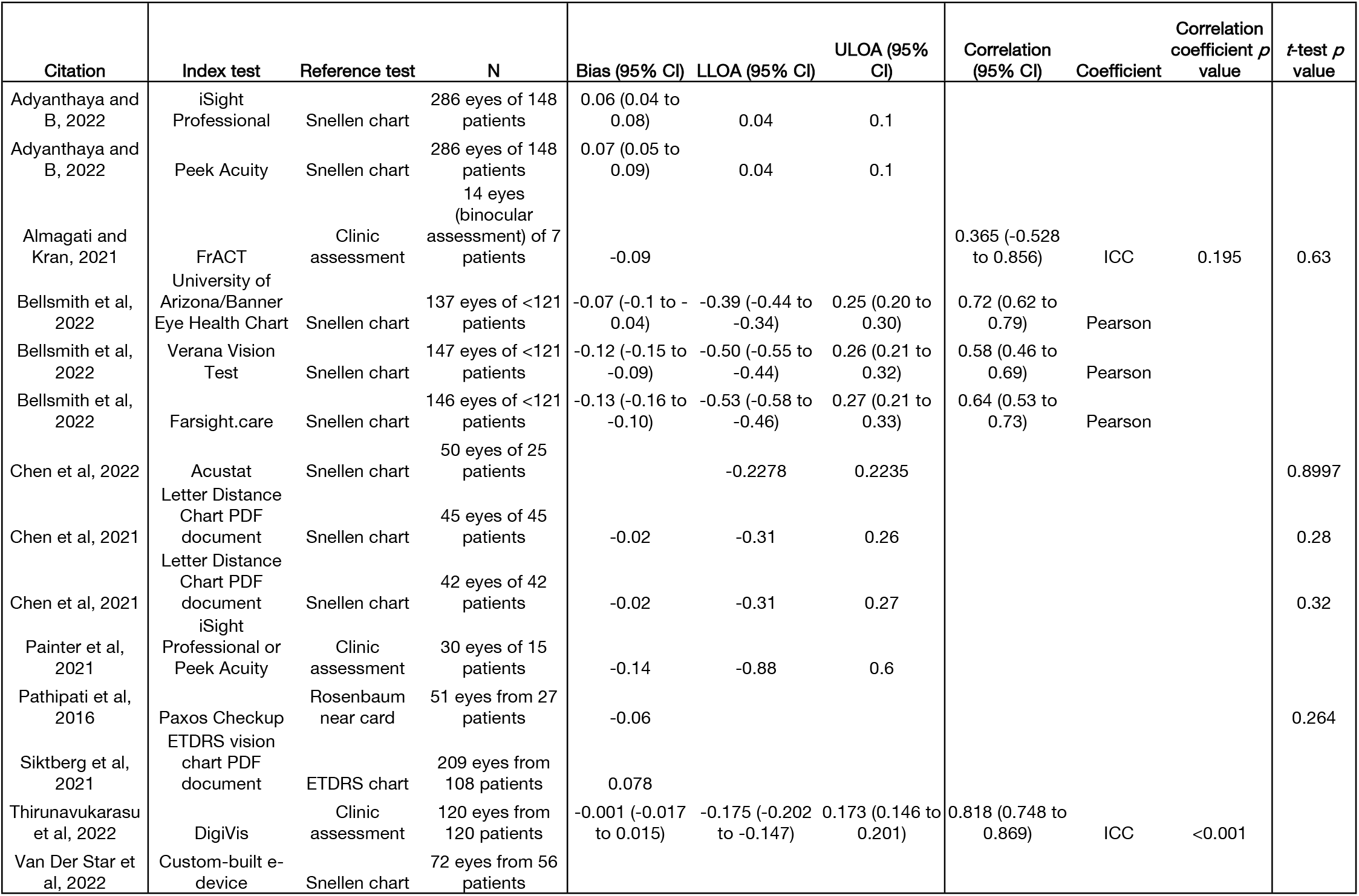
Accuracy data. Comparing remote index tests to clinical reference tests. CI = confidence interval; LLOA = lower 95% limit of agreement; ULOA = upper 95% limit of agreement; ICC = intraclass correlation coefficient; PDF = portable document format; ETDRS = Early Treatment of Diabetic Retinopathy Study.

Eight studies provided Bland-Altman statistics, corresponding to trials of twelve remote VA tests (Figure 3).^17,19–25^ Of these, six studies (ten trials) provided 95% lower and upper limits of agreement (LLOA and ULOA respectively).^17,19–22,25^ Three trials’ LOA lay within ±0.2 logMAR, corresponding to Isight pro, Peek Acuity, and DigiVis.^17,25^ The remaining seven trials corresponded to University of Arizona/Banner Eye Health Chart, Verna Vision Test, Farsight.care, Acustat, Letter Distance Chart PDF document (twice), and Isight pro or Peek Acuity pooled.^19–22^ One study did not report the bias; of the remaining nine studies, three (containing six trials) provided 95% confidence intervals.^17,19,25^ Isight pro and Peek Acuity exhibited significantly higher bias than 0 logMAR (index test estimated worse acuity);^17^ University of Arizona/Banner Eye Health Chart, Verana Vision Test, and Farsight.care exhibited significantly lower bias than 0 logMAR (index test estimated better acuity);^19^ and DigiVis exhibited no statistically significant bias.^25^ The mean magnitude of the bias was 0.04 logMAR (2 letters).^17–19,21–25^ Three studies (5 trials) reported (or facilitated calculation of) correlations: Pearson coefficients ranging from 0.58 to 0.72,^19^ and two ICCs ranging from 0.365 to 0.818.^18,25^ Four studies’ (five trials) *t*-tests comparing measurement methods all reported *p*-values above 0.25.^18,20,21,23^

**Figure 3.**
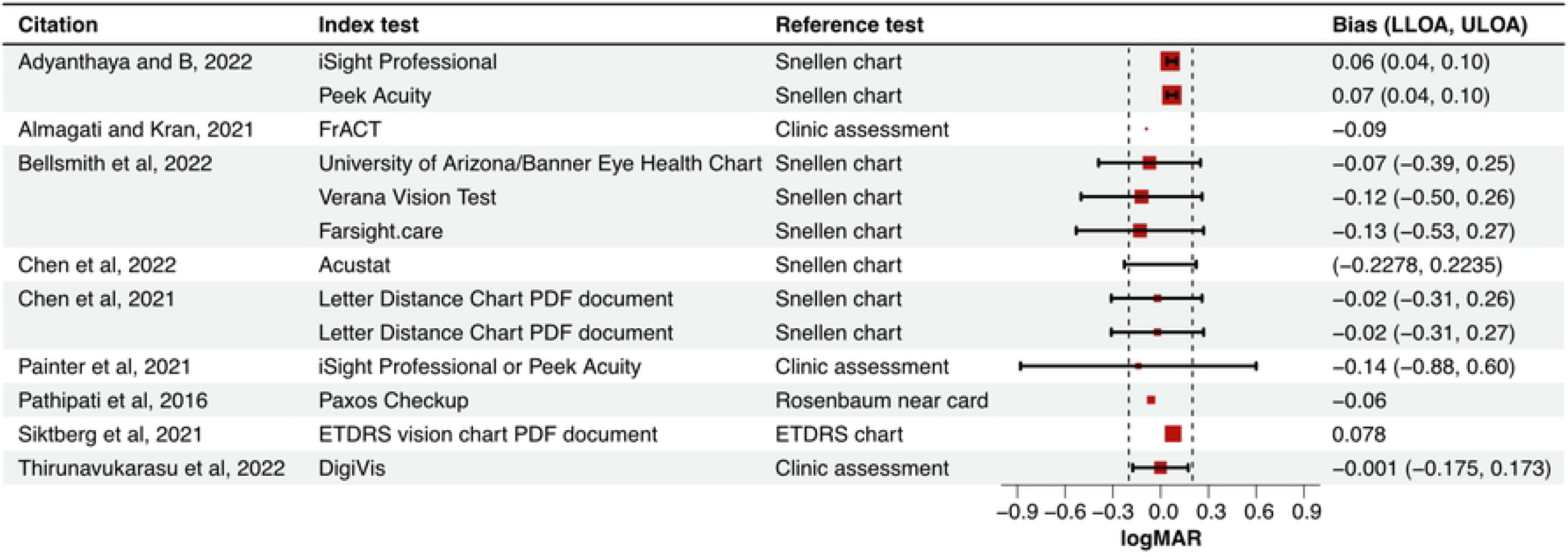
Forest plot summarising Bland-Altman analyses of accuracy. LLOA = lower 95% limit of agreement; ULOA = upper 95% limit of agreement; PDF = portable document format; ETDRS = Early Treatment of Diabetic Retinopathy Study; logMAR = logarithm of the minimum angle of resolution.

Two trials reported test-retest reliability: one trialling DigiVis,^25^ and one trialling Isight pro and Peek Acuity in a pooled analysis.^22^ The former reported Bland-Altman statistics and intraclass correlation coefficient, whereas the latter only reported the coefficient of repeatability (Table 3). DigiVis exhibited a bias equivalent to 0, LOA of ±0.12 logMAR (6 letters), and ICC of 0.922.^25^ In a pooled analysis, Isight pro and Peek Acuity exhibited a coefficient of repeatability of 0.03 logMAR.^22^

**Table 3.** Test-retest agreement. Assessing the reliability of remote tests. CI = confidence interval; LLOA = lower 95% limit of agreement; ULOA = upper 95% limit of agreement; CoR = coefficient of repeatability; ICC = intraclass correlation coefficient.

| Citation | Test | N | Bias (95% CI) | LLOA (95% CI) | ULOA (95% CI) | CoR (95% CI) | Correlation (95% CI) | Correlation Coefficient | Correlation coefficient p value |
| --- | --- | --- | --- | --- | --- | --- | --- | --- | --- |
| Painter et al, 2021 | iSight Professional or Peek Acuity | 26 eyes of 13 patients |  |  |  | 0.03 (-0.08 to 0.04) |  |  |  |
| Thirunavukarasu et al, 2022 | DigiVis | 105 eyes from 105 patients | 0.001 (-0.011 to 0.013) | -0.121 (-0.142 to -0.101) | 0.124 (0.103 to 0.144) |  | 0.922 (0.887 to 0.946) | ICC | <0.001 |

## Discussion

To justify adoption of remote self-administered VA tests, there must be convincing evidence that the proposed platform meets regulatory safety standards, is effective enough to fulfil its clinical function, is accessible to patients—with appropriate mechanisms to serve those unable to use the platform, and is economically viable.^38^ Facilities for VA self-assessment may be useful in a number of domains: improving the capacity and capability of teleophthalmology clinics, empowering patients with the ability to monitor their own vision rather than attend regular appointments; enabling non-eye specialists to obtain useful information for a referral to ophthalmology; and giving eye units a tool to facilitate pre-attendance triage of eye casualty cases.^2,3^ In all cases, it is essential that tests are accurate and reliable, exhibiting agreement with clinical assessment and with repeated remote measurement, respectively.

In ideal conditions, chart-based VA still exhibits considerable variation, with 95% LOA approaching 0.09 logMAR.^10^ In reality, clinical variation is greater as different examinations may be more or less demanding of patient effort, and may or may not test to majority failure (*i*.*e*. ≥3 errors on 1 line).^39^ Where both index and reference test exhibit variation, the utility of analyses restricted to *t*-tests or correlation coefficients is limited. Bland-Altman analysis compensates for bivariate variation by quantifying 95% LOA, which provides metrics of measurement dispersal which can be compared to gold-standard tests. Studies failing to conduct appropriate analyses fail to provide evidence of validation—it is not possible to ascertain whether observed variation is clinically acceptable or not. Acceptable 95% LOA should compare well with those exhibited by conventional clinical chart-based tests: below ±0.2 logMAR.^39^ Bias should be close to zero—statistically significant deviation (*e*.*g*. if confidence intervals do not cross zero) indicates a systematic error. High correlation is expected—over 0.7 in terms of Pearson’s or intraclass correlation coefficients.^40,41^

Here, DigiVis was the only test exhibiting undisputed 95% LOA within 0.2 logMAR, no significant bias, and high correlation between remotely and clinically assessed VA.^25^ iSight Professional and Peek Acuity exhibited 95% LOA within 0.2 logMAR in one of two studies, but this study was judged to be at a high risk of bias.^17^ In the trial finding greater LOA, pooling of results from both tests may have affected calculated accuracy.^22^ Just two studies reported test-retest agreement. One study indicated that DigiVis measurements are very reliable;^25^ while another indicated good agreement between repeated iSight Professional and Peek Acuity measurements, albeit with few statistics provided.^22^ Again, pooling of iSight Professional and Peek Acuity data may have affected the result.^22^

All three tests with positive validation data had no requirement for real-time administration by a trained clinician. Therefore, all three may be used to improve the capability of telehealth services and eye assessment by non-specialists such as general practitioners and emergency department clinicians. However, as some patients in the DigiVis trial conducted the remote test in clinical settings, it is difficult to conclude with certainty that deployment for home-based assessment is justified.^25^ All three tests relied on digital devices, accessible by most of the world’s population.^42^ As uptake of smartphone-based vision tests correlates negatively with older age and worse vision, healthcare providers should be mindful of patients’ capacity to access and complete remote VA assessment to ensure their care and outcomes are not adversely affected.^37^

This review was limited by three factors: (1) Inconsistent and incomplete statistical analysis made establishing the accuracy and reliability of trialled VA tests challenging. *De novo* analysis and interpolation were conducted where amenable data were provided, and conclusions were restricted to what was indicated by available data. Deduction of the direction of bias was often based on limited prose descriptions—this is a potential source of error but would not affect conclusions significantly as bias was always close to 0. (2) Descriptions of the setting of the remote index test was often unclear, making the full-text screening process more difficult. Included studies all mentioned a test undertaken outside the eye clinic and did not state that all tests were conducted in clinical or ideal settings. (3) Most studies did not use Bailey-Lovie or ETDRS charts which are accepted as more accurate and precise for clinical research. While this may inflate variability in the reference test and consequently inflate calculated accuracy of the remote index tests, use of Snellen chart may not be a specific weakness as it remains widespread in clinics around the world.^43,44^

Although promising technology has been developed to remotely assess VA, very few studies have demonstrated that patient-led assessment outside the eye clinic is feasible. DigiVis, iSight Professional, and Peek Acuity all have validation data demonstrating equivalence with clinical assessment, with the former being best justified due to conflicting results regarding the latter two tests. Further pragmatic trials are required to demonstrate the accuracy and reliability of remote VA assessment to justify deployment at scale—ongoing preregistered trials may fill this significant gap in the literature base.^45,46^ However, as these trials are organised by test manufacturers, owners, or patent-holders, external research teams may seek to run their own studies to ensure validation data are unbiased. Reporting must be comprehensive—particularly in descriptions of patient characteristics, index test setting, and statistical analysis. Validated self-administered VA tests have the potential to augment teleophthalmology services, pre-consultation triage, long-term monitoring, as well as non-specialist assessment and reporting of eye problems.^3^

## Data Availability

Data will be published upon acceptance.

## Legends

**S1 Figure. Summarised risk of bias and inapplicability**. Appraised with the QUADAS-2 framework. QUADAS-2 = Quality Assessment of Diagnostic Accuracy Studies 2; RoB = risk of bias; CrA = concerns regarding applicability.

